# An Exploration of the Mental Health impact among Menopausal Women: The MARIE Project Protocol (UK arm)

**DOI:** 10.1101/2023.11.06.23298158

**Authors:** Gayathri Delanerolle, Anna K Forbes, Heitor Cavalini, Julie Taylor, Kathleen Riach, Sharron Hinchliff, Carol Atkinson, Kristina Potocnik, Paula Briggs, Om Kurmi, Vikram Talaulikar, Jeremy Van Vlymen, Ashish Shetty, Muhammed Irfan, Rabia Kareem, Helen Kemp, Sanghamitra Pati, Subrata Kumar Palo, Nirmala Rathnayake, Lucky Saraswat, Toh Teck Hock, Jian Qing Shi, Peter Phiri

## Abstract

**Introduction:** Menopause marks the end of the menstruation period which can incur naturally or due to surgery where the ovaries or the uterus is removed, or the use of other treatments like chemotherapy. Menopause elicits both physiological and psychological changes such as joint or pelvic pain, headaches or migraine, cognitive function and mental health problems such as anxiety. In order to assess the mental health impact of menopause, the physiological, psychological and sociological composites need to be evaluated. It is increasingly recognised that the associated symptoms experienced by women and trans-men are specific to menopause transition, making it challenging to diagnose and treat using conventional methods. We developed a menopause tool called MenopAuse mental hEalth (MARiE) rating tool following a co-production workshop. The MenopAuse mental hEalth (MARIE) project’s overall aim is to explore the mental health impact of menopause through several work stream packages and assess the MARiE tool. The current work package (WP 2a and 2b) that is represented within this study aim to further explore menopause symptoms and then validate and, determine the efficacy of the MARiE tool.

**Methods:** We will conduct a prospective mixed methods study in the United Kingdom (UK) among women and trans-men ≥18 years old that are perimenopausal, menopausal or post-menopausal. The quantitative portion will use the Hospital Anxiety and Depression Scale, Insomnia Severity Index Scale, Menopause Rating Scale, Greene Climacteric Scale, Health related quality of life, Quebec Pain Disability Scale, and Burnout Assessment Tool in addition to the MARiE tool within the scope of WP2a and WP2b, respectively. The qualitative component will use a topics guide.

Research Ethics and Health Research Authority approval has been obtained with a reference of 22/EE/0159.

**Dissemination:** The findings will be submitted for publication in peer-reviewed journals and presented at women’s health, primary care, and mental health themed conferences.

## INTRODUCTION

The menopause transition is defined as the time-period from the start of changes to the menstrual cycle or vasomotor symptoms, until 12 months after the final menstrual period^1^. The final menstrual period is termed the menopause. Menopause transition is marked by hormonal changes and is associated with a various of physical and psychological health problems, which vary substantially in both nature and severity across different geographical locations and cultures^2^. Typically, women first experience symptoms in their mid to late 40s and they persist for 5 to 8 years, but may continue long term^3^.

Women have a two-fold higher risk of developing depression or anxiety disorders than men^4^. Changes in cognitive function and mental health problems are commonly experienced during menopause transition^5^. However, few dedicated tools exist to explore and manage these important issues.

The Nuffield Health Survey of 3725 menopausal women in the UK found that 47% reported symptoms of depression and one third experienced feelings of anxiety^6^. Moreover, sleep disturbance, including difficulty falling asleep and repeated awakenings is experienced by up to one third of women during menopause transition, and can exacerbate mental health issues^7^. It is approximated that up to 20% of women in menopause transition attend their primary care physician with symptoms of depression or anxiety^8^, illustrating the gravity and impact of the problem.

Findings from the Study of Women’s Health Across the Nation (SWAN) established that numerous factors contribute to the susceptibility to depression during this time. This includes discomfort associated with physical symptoms such as hot flushes, pre-existing mental health conditions, insufficient social support, psycho-social stressors, health and lifestyle behaviours, and demographic attributes^5^. However, the increased risk of depression and anxiety exhibited in this population is not fully explained by these factors. Evidence from studies adjusting for these variables demonstrates a residual increased risk^9,10^.

It is increasingly recognised that the clinical presentation of depression and anxiety is different in women during menopause transition compared to women not undergoing these biological changes. It is associated with a broad range of symptoms including fatigue, lack of energy, low self-esteem, feelings of loneliness, cognitive impairment, reduced libido, weight gain, muscle aches and back pain^11^. The mood changes experienced by women during menopause transition are predominantly irritability, paranoia, and anger, compared with women of reproductive age who typically display sadness and low mood^12^.

The aetiology of mental health issues during menopause transition is multi-dimensional and includes biological, psychological, and social influences. Hormonal changes in the hypothalamic-pituitary-gonadal axis have a direct impact of brain functioning. Gonadal hormones play and important role in the regulation of brain metabolism, increasing cerebral blood flow, reducing inflammation, and simulating neuronal regeneration and nerve growth factor^13 14^. These biological influences combine with psycho-social factors occurring at this life period, including the perception of ageing, the end of reproductive capabilities, changing roles within the workplace and family, physical illness, and stressful life events.

The evidence supports the view that mental illness during this period has a unique aetiology and specific clinical features^15^, highlighting the need for a dedicated tool to explore and evaluate mental health problems during menopause transition.

## METHODS

### Aims and Objectives

This project aims to explore the mental health impact of menopause by way of evaluating the physiological and psychological aspects through several work stream packages. This protocol focusses on work package 2 which comprises of two parts: work packages 2a and 2b. The overall objective of work package 2 is to better explore menopause in a variety of populations and further validate of our new menopause tool known as the MenopAuse mental hEalth Rating (MARiE). The tool was constructed following an evidence synthesis of existing research data, clinical expertise and a co-production piece conducted as part of a wider women’s health program; ELEMI.

### Study Design

WP2a is a prospective mixed methods study conducted to gather further information using clinically validated questionnaires that are currently in use and a study specific topics guide. Participants will be required to complete a brief online demographic questionnaire followed by core questions about menopause symptoms, mental health and wellbeing using Hospital Anxiety and Depression Scale (HADS), Insomnia Severity Index Scale (ISIS), Menopause Rating Scale (MRS), Greene Climacteric Scale (GCS), Health related quality of life (HRQoL), Quebec Pain Disability Scale (QPDS), and the Burnout Assessment Tool (BAT-12). The questionnaires will be completed online using the Qualtrex tool on day 1 and 30, and those that consent will additionally participate in a qualitative interview on day 60. Some of the questionnaires are generic and are part of clinical consultations as part of the current healthcare landscape. However, their non-specificity to the context of menopause can limit its’ purpose to better understand experiences of those experiencing the menopause. However, it is recognised some questions may still have validity and these will be repurposed as part of the MARiE tool.

Work package 2b is a prospective mixed methods study on validating the MARiE assessment tool in a winder population of participants with a view to demonstrate its’ feasibility and acceptability.

### Study population & recruitment

The eligibility criteria includes women and trans-men ≥18 years of age that have undergone or undergoing natural or surgical menopause, peri-menopause and post-menopause. We will also include trans-men who are using hormone replacement therapy. We aim to recruit at least 100 people for WP2a and WP2b. Participants must be willing and able to give electronic consent and have access to a digital device to complete the online questionnaires and interviews. Participants will be invited to participate via social media platforms such as Twitter, LinkedIn, National Health Service (NHS) hospital websites, Instagram and Facebook.

### Informed consent

Informed consent will be given electronically using the Qualtrics XM platform, and separately for each component of work package 2. Participants will be given detailed information about the nature and purpose of the study and will be required to confirm that they meet the inclusion criteria. Eligible individuals that have given consent will then be enrolled into the study and asked to complete the relevant online questionnaires on day 1 and day 30. Additionally, participants will be invited to provide their email or telephone contact details if they are happy to take part in a qualitative exploration via interviews. Participants will also have an option to consent to be contacted for future research.

Participants will have the opportunity to withdraw from the study prior to submitting their questionnaire. However, once submitted we will not be able to identify the participant to withdraw their data, unless they have provided contact details which could be used to identify and withdraw them. Under these circumstances data already collected with consent would be retained but no further data would be collected, or any other research procedures carried out in relation to the participant.

### Data collection

Quantitative data will be collected through online questionnaires using the Qualtrics XM platform. Participants will be allowed to omit questions they do not want to complete. All participants will be allocated a study ID and patient identifiable information will not be recorded. Survey data will be exported from the Qualtrics XM platform to the statistical software packages SPSS and STATA for graphical presentation and analysis.

Topic guides have been developed to facilitate the participant interviews. All interviews will be conducted via a secure online facility, audio-recorded, transcribed in full, and early interviews will be reviewed by the research team to determine whether any alterations to the topic guides need to be made. Data collection and analysis will be integrated. If data saturation is reached, interview capacity will be shifted to other groups to facilitate greater exploration where appropriate. It is anticipated that interviews will take approximately 45 minutes, however this will be guided by each participant.

Interviews will be transcribed by the researchers. Transcripts will then be reviewed for accuracy, de-identified, and uploaded to NVivo software version 11.2 for data coding and retrieval.

### Outcome measures

The primary objective of work package 2a is to determine the mental health impact of the menopause using established tools. The outcome measures are the responses to the questionnaires, including the HADS and ISIS, in addition to participant interviews using topic guides.

The secondary objective is to explore other effects of the menopause. The outcome measures are the responses to the questionnaires from the MRS, GCS, HRQoL, QPDS, and BAT-12. We will evaluate challenges associated with the menopause, specifically lower back pain, vasomotor symptoms, cognitive performance, and wellbeing at work.

The primary objective of work package 2b is to determine the validity and effectiveness of the MARiE tool. The outcome measures are the responses to the completed MARiE questionnaires, in addition to participant interviews using topic guides.

### Statistical analysis

WP2a: Descriptive statistics will be presented as either means (SD) or median (IQR) for continuous variables according to the distribution of data. Descriptive statistics for categorical data will be presented as frequencies and proportions. Where data is not normally distributed, non-parametric techniques (Kruskal-Wallis test and Mann-Whitney U-test) will be utilised. Chi-square or Fisher’s exact test will be used for categorical variables and any associations between demographic data and responses related to mental health wellbeing questions. Pearson’s correlation analysis will be used to measure strength of linear relationship between variables.

WP2b: Pearson’s correlation analysis will be used to measure strength of linear relationship between variables. The goodness-of-fit of latent class models for categorical responses can be tested using Pearson and likelihood-ratio chi-squared tests. This will be followed by a factor model, with confirmatory factor analysis using the Maximum likelihood approach. The data’s covariance fit for the tested factor models will be explored. Models will be used based on their goodness of fit-indices (GOF). Varies indices, absolute fit or relative fit, will be considered.

All interviews will be conducted via a secure online facility (password protected zoom teleconference), and will be audio-recorded and transcribed in full. Early interviews will be reviewed by the research team to determine whether any alterations to the topic guides need to be made. Data collection and analysis will be integrated. A process based on framework methodology will be used to analyse the data using an indexing coding approach to charter the data to facilitate the interpretation.

## Data Availability

All data produced in the present study are available upon reasonable request to the authors

## AUTHOR’S CONTRIBUTIONS

GD conceptualised and developed the MARiE project as part of the ELEMI program. GD and JQS designed the statistical analysis plan. GD, PP, AF and HC drafted the protocol manuscript. All authors critically appraised and commented on the protocol manuscript. All authors read and approved the final manuscript.

## DATA AVAILABILITY STATEMENT

The authors will consider sharing the dataset gathered upon receipt of reasonable requests.

## List of abbreviations

MARiE: Menopause mental health Rating
GOF: Goodness of Fit

